# Use of Drama for Improving Breastfeeding Initiation, Exclusive Breastfeeding and Breastfeeding Self-efficacy among Rural Pregnant Women from Selected Communities in Two Local Government Areas (LGAs) in Ibadan, Nigeria

**DOI:** 10.1101/2023.08.03.23293594

**Authors:** Ogundairo Omotola Yetunde, Adepoju Oladejo Thomas, Olumide Olufunmilola Adesola

## Abstract

**Background:** Breastfeeding self-efficacy (BFSE) is a key variable that enhances exclusive breastfeeding (EBF) and promotes positive health outcomes for infants and their mothers. To increase BFSE and EBF of mothers, numerous interventions targeting prenatal and postnatal periods have been developed. However, there is paucity of studies utilizing drama interventions for improving BFSE and EBF.

**Objectives:** This study assessed the effect of drama usage on breastfeeding self-efficacy, initiation, and exclusive breastfeeding of pregnant women in rural communities in Lagelu and Egbeda Local Government Areas (LGAs).

**Methodology:** A quasi-experimental designed was carried out with pregnant women in their second trimester. Selected communities from Lagelu and Egbeda LGAs were randomized into experimental and control groups. A total of 200 pregnant women (100 experimental and 100 control groups) were enlisted as well as followed-up at one, three, and six months after giving birth. Six-session program comprising four episodes of drama and two sessions on hygiene practices were presented to experimental groups in the communities prior to delivery, while the control group received only hygiene talk. An electronic questionnaire (ODK) was used to obtain information on socio-demographic, BFSE, initiation, and exclusive breastfeeding during prenatal and postnatal periods. BSFE score was categorized as low (14–32), average (33–51), and high (52–70). Descriptive and inferential statistics was used to analyzed data α0.05.

**Results:** The experimental and control mean ages were 28.40 ±6.50 and 27.00±6.20 respectively. Average and high BSE pre-intervention (11.0%; 89.0% and 9.0%; 91.0%) and six months post-intervention (97.3%;100% and 95.2%; 95%) for intervention and control. Age, marital status and occupation were predictors of BFSE (R^2^ = 22.3; p<0.001)

**Conclusion:** Experimental women had an increase in BFSE, initiation, and EBF practice compared to control women.

## INTRODUCTION

Breastfeeding self-efficacy enhances a woman’s ability to confidently breastfeed her infants. It predicts the mother’s decision on breastfeeding, the preferred method she plans to use in feeding her infants, how much effort she will use to achieve this, her thought pattern, as well as her emotional reactions to breastfeeding difficulties (1). Several studies have identified the linkage between breastfeeding self-efficacy, breastfeeding intention, initiation, duration, and exclusivity (2–3). However, breastfeeding self-efficacy was identified as a key factor that influences maternal perceptions and behaviors (4–5) and this has is also associated with maternal depression, postpartum depression, perceived insufficient breastmilk, and social support (6,7).

The four drivers of breastfeeding self-efficacy used to sustain an individual health behavior was used to develop a breastfeeding self-efficacy intervention model to increase the confidence of mothers to breastfeed their infants (4). This model increased the confidence of mothers to breastfeed their infants at home, in the community, and in hospitals. During the prenatal and postnatal periods, different educational material such as flip charts, pamphlets, videotapes, skin-to-skin contact, and health talk have been used to increase the breastfeeding self-efficacy of mothers (8–10). The findings showed a positive increase in breastfeeding self-efficacy as well as outcomes at one month, three months, and six months postpartum, respectively.

Little is known about the use of drama intervention to improve breastfeeding self-efficacy, initiation, and exclusive breastfeeding of mothers. Drama, which has a performance and interdisciplinary attribute for multiple representations and understanding of important questions as well as ensuring the synergy between cognitive and emotional domains, has been used in the health sector, but little has been documented on its use in the promotion of breastfeeding. Van de Water (2021) noted that research from other disciplines such as sociology, neuroscience, and psychology has started to embrace the position of drama practitioners that “human instincts call for embodied and contextualized learning”. Drama not only encourages participants and viewers to envision possibilities in their thoughts but also creates opportunities for them to express abstractly conceived ideas (11). There is still death of information on the use of drama for improving breastfeeding self-efficacy, breastfeeding initiation, and exclusive breastfeeding of women. This study was therefore designed to assess the use of drama intervention to improve breastfeeding self-efficacy, initiation, and exclusive breastfeeding of women from selected communities in Ibadan, Oyo State, Nigeria.

## METHODOLOGY

### Study design and participants

A quasi-experimental study was conducted over a period of 18 months (June 2020 to December 2021). A total of 200 pregnant women in their second trimester who gave their voluntary consent were recruited from selected rural communities in Egbeda and Lagelu Local Government Areas and randomized into the intervention (n=100) and control groups (n=100). A pre-tested, self-administered questionnaire consisting of five sections was used to obtain information. Each section includes: maternal socio-demographic characteristics, pregnancy related issues (pregnancy trimesters, parity, mode of delivery, antenatal registration, and sources of care, breastfeeding self-efficacy, breastfeeding initiation, and exclusive breastfeeding practices. Breastfeeding self-efficacy was assessed using with breastfeeding self-efficacy short form tools (BSES-sf), 14-item scale revised by Dennis (2003). This was initially a 33-item scale by Dennis and Faux (1999). Breastfeeding initiation and exclusive breastfeeding was assessed using the breastfeeding practices questions by Agboh et al., (2007). A total of 14 questions were asked from the respondents. Each question was graded on a five-point Likert scale: 5 = Very Confident, 4 = Confident, 3= Sometimes confident, 2 = not very confident, and 1 = not at all confident. The total score from each respondent was categorized as low self-efficacy (14–32), average self-efficacy (33–51) and high self-efficacy (52–70) (14–15).

### Data collection

A computer aided personal assistant (open data kit) was used to obtain information from respondents at baseline and endline (one, three and six month postpartum). At baseline, information on socio-demographic characteristics, pregnancy-related issues, and breastfeeding self-efficacy was obtained. Also at baseline, a breastfeeding chart containing the expected date of delivery, names, and phone numbers of the participants and their spouses was developed to monitor each respondent delivery days for other follow-up data. Peer-support women who also lived in the various communities assisted in monitoring the participants and provided daily delivery reports on the time and day of delivery. After they were discharged from the hospital, follow-up data on breastfeeding initiation time, exclusive breastfeeding practices, and breastfeeding self-efficacy was also obtained at their homes.

The first follow-up was conducted within one week postpartum for the participants who gave birth to obtain information on their breastfeeding initiation time. At one month postpartum, the second follow-up was conducted to assess their confidence on exclusive breastfeeding practice. At three and six months postpartum, The third follow-up and fourth follow-up was conducted at to also assess the mother’s confident and practice of exclusive breastfeeding. Participants in the control group received a two-episode, step-by-step practical session on hand washing and personal hygiene.

A drama-based intervention model was designed with the four drivers of breastfeeding self-efficacy to increase the breastfeeding self-efficacy of participants in the intervention group prior to their delivery. They viewed a recorded 80-minute, four-episode drama-based intervention in open village squares. This was done concurrently in the two LGAs every fortnight for a period of two weeks. The recorded drama piece contains information on early initiation and importance of colostrum, proper positioning and attachments, perceived insufficient breast milk and breast size, advantages of breastfeeding to mothers and infants, and mothers past and present successes were all covered in the recorded drama. For the first week, the women viewed two episodes of the drama containing information on early initiation, the importance of colostrum, proper positioning and attachments in breastfeeding, and perceived insufficient milk for a period of 40 minutes. The second episode contained information on the breast size and quantity of breastmilk, the benefits of breastfeeding to mother, child, and community, and mothers past and present experiences with breastfeeding. At the end of each episode, there was a brief discussion with the participants to assess their level of understanding and knowledge, and this was used as a basis for further understanding in the next episode.

### Ethical approval

Ethical approval was obtained from the University College Hospital and University of Ibadan Ethical Review Committee and Oyo State Ethical Review Committee

### Data Analysis

Baseline data were summarized with descriptive statistics. Independent t-test was used to compare mean values, Chi-square test was used to test for association between variables, repeated measure of Analysis of variance (ANOVA) was used to determine change in breastfeeding self-efficacy mean scores, and logistic regression was used to identify other predictors of breastfeeding self-efficacy at p<0.01.

## RESULTS

Table 1 explains the socio-demographic characteristics of women in the study group. The overall mean ages of the respondent were 27.00±6.20 and 28.40±6.50 for intervention and control groups. Slightly above half (53.5%) were between 20 – 29 years of age, and 52.5% were Christians. Almost all (97.0%) participants were married, 90.5% were Yoruba, and 60% had a secondary level of education as their highest level of education. Most (89.5%) participants earned less than the government-regulated monthly minimum wage of ₦30,000.00, and 37.5% were traders. There was no significant difference in the socio-demographic characteristics of the participants in the intervention and control groups (p>0.01).

**Table 1:**
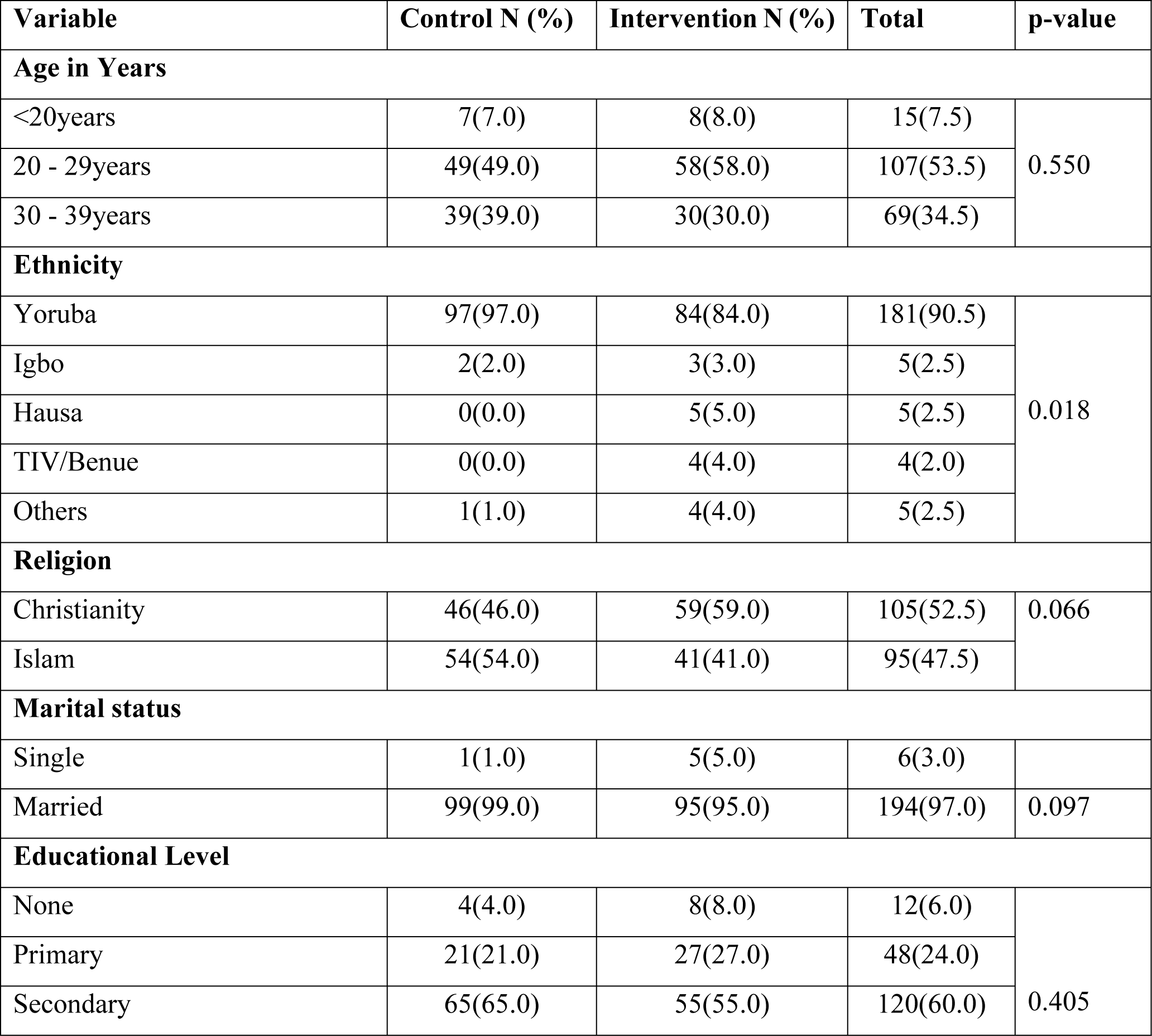
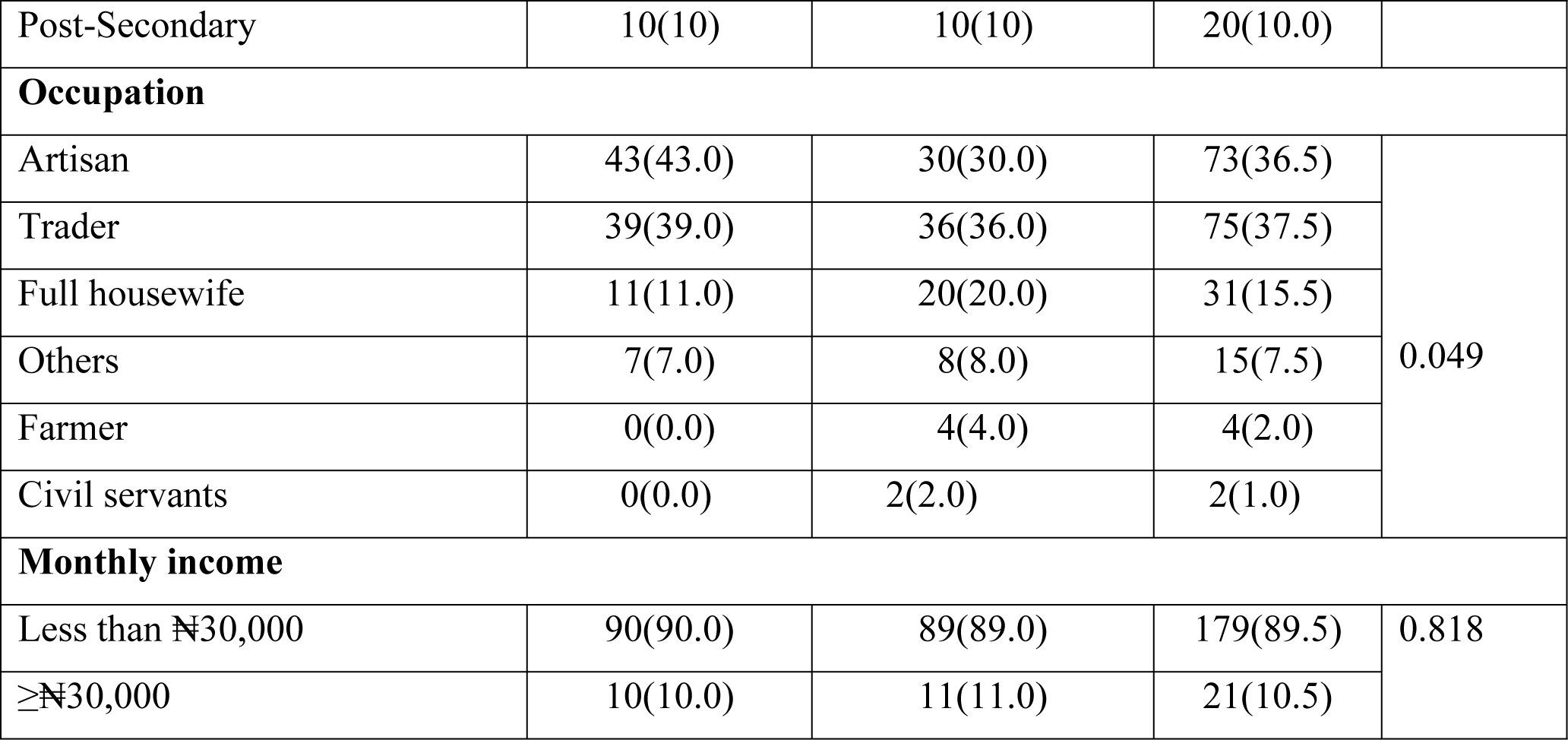
Socio-demographic characteristics of participants.

In Table 2, 49.0% of the participants were in their second trimesters of pregnancy, 77% were multiparous mothers, 73.5% had registered at the antenatal clinic, and 76.0% gave birth to their previous child through normal spontaneous vaginal delivery.

**Table 2:**
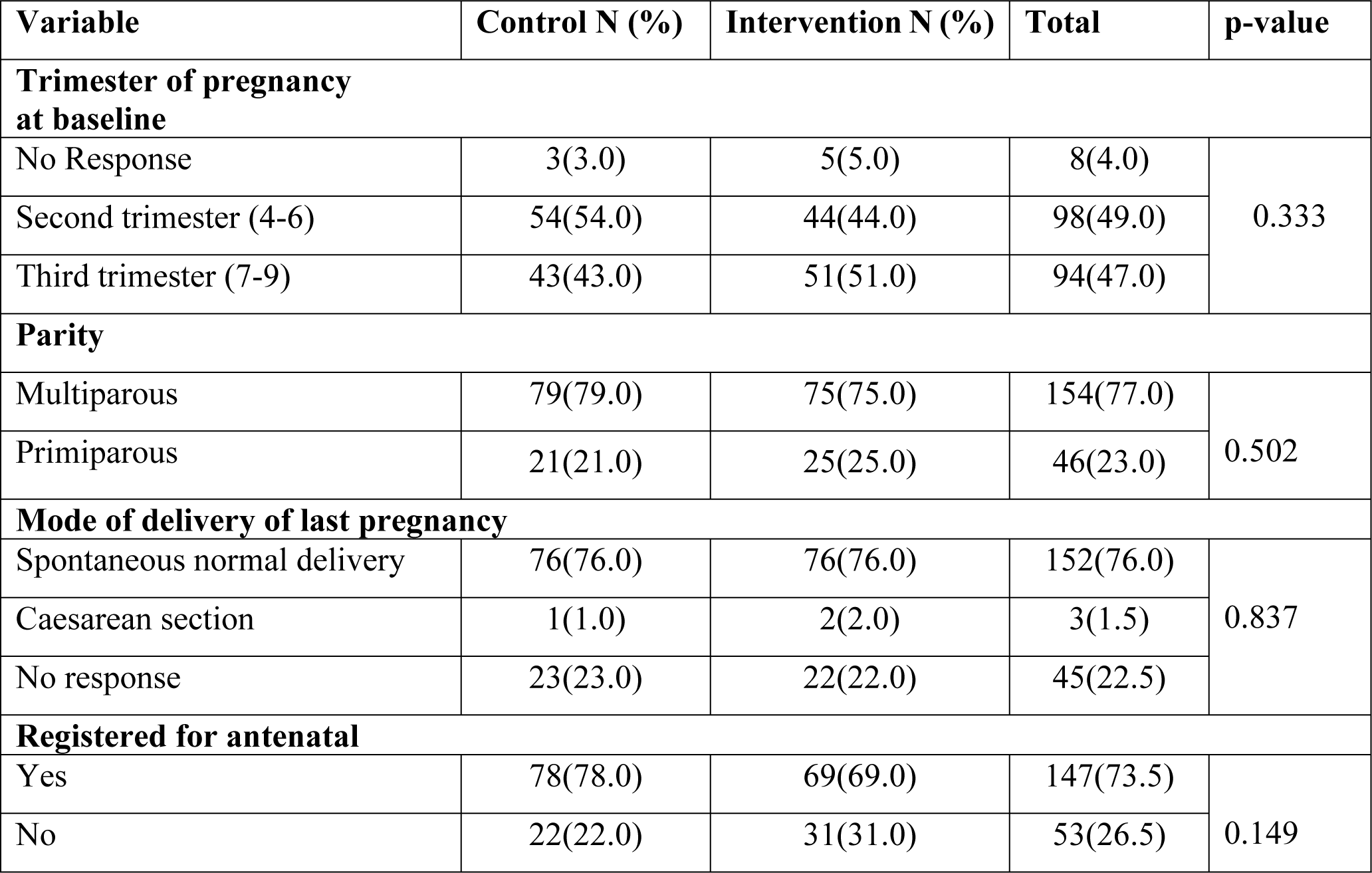
Pregnancy history of participants.

| Variable | Control N (%) | Intervention N (%) | Total | p-value |
| --- | --- | --- | --- | --- |
| Trimester of pregnancy at baseline |  |  |  |  |
| No Response | 3(3.0) | 5(5.0) | 8(4.0) | 0.333 |
| Second trimester (4-6) | 54(54.0) | 44(44.0) | 98(49.0) |  |
| Third trimester (7-9) | 43(43.0) | 51(51.0) | 94(47.0) |  |
| Parity |  |  |  |  |
| Multiparous | 79(79.0) | 75(75.0) | 154(77.0) | 0.502 |
| Primiparous | 21(21.0) | 25(25.0) | 46(23.0) |  |
| Mode of delivery of last pregnancy |  |  |  |  |
| Spontaneous normal delivery | 76(76.0) | 76(76.0) | 152(76.0) | 0.837 |
| Caesarean section | 1(1.0) | 2(2.0) | 3(1.5) |  |
| No response | 23(23.0) | 22(22.0) | 45(22.5) |  |
| Registered for antenatal |  |  |  |  |
| Yes | 78(78.0) | 69(69.0) | 147(73.5) | 0.149 |
| No | 22(22.0) | 31(31.0) | 53(26.5) |  |

**Table 3:** Regression analysis.

| Model | B | SE | T | Sig | 95.0% Confidence Interval for B |  |
| --- | --- | --- | --- | --- | --- | --- |
|  |  |  |  |  | Lower bound | upper bound |
| Age | 0.239 | 4.935 | 2.580 | 0.11 | 0.56 | 0.422 |
| Marital Status | 6.379 | 3.235 | 2.083 | 0.39 | 0.335 | 13.142 |
| Occupation | 3.067 | 1.406 | 2.182 | 0.31 | 0.284 | 5.849 |

**Table 4:** Summary of Model analysis.

| Model<br>summary | R | R Squares | Adjusted R square | Standard error of estimate | F | Sig |
| --- | --- | --- | --- | --- | --- | --- |
|  | 0.472 | 0.223 | 0.108 | 5.02845 | 1.946 | 0.018 |

### Breastfeeding initiation of women by study group

In Figure 1, 52.3% of the participants in the control group initiated breastfeeding within the WHO recommended period of the first hour of birth, while 65.5% initiated breastfeeding within the WHO recommended standard of one hour in the intervention group, showing that there was a significant difference between the proportion of women who initiated breastfeeding early in the intervention group compared with the control group (p<0.01).

**Figure 1:**
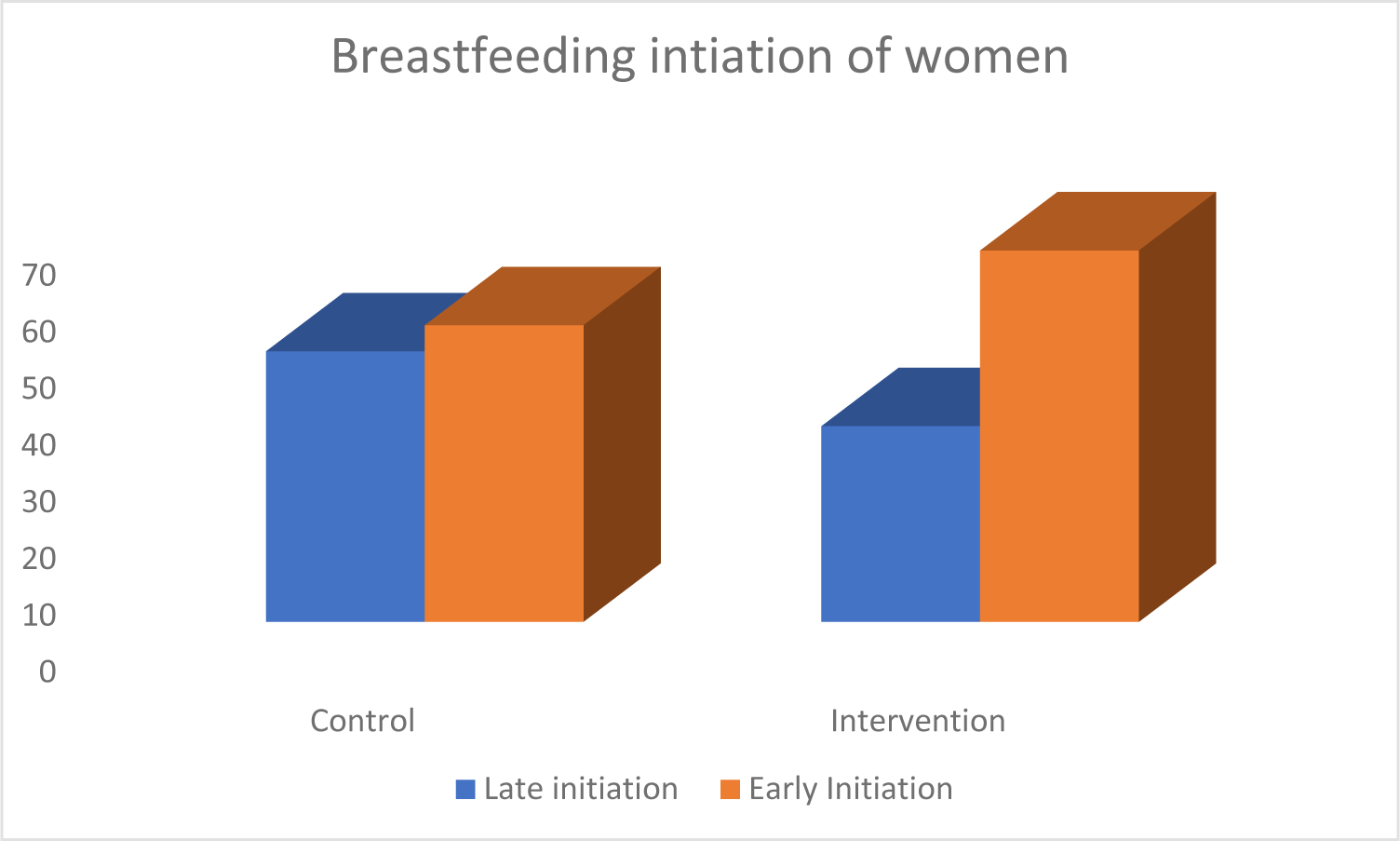
Breastfeeding initiation of women

In Figure 2, 22.9% of participants in the control group practiced exclusive breastfeeding (EBF), while 43.2% practiced EBF in the intervention group.

**Figure 2:**
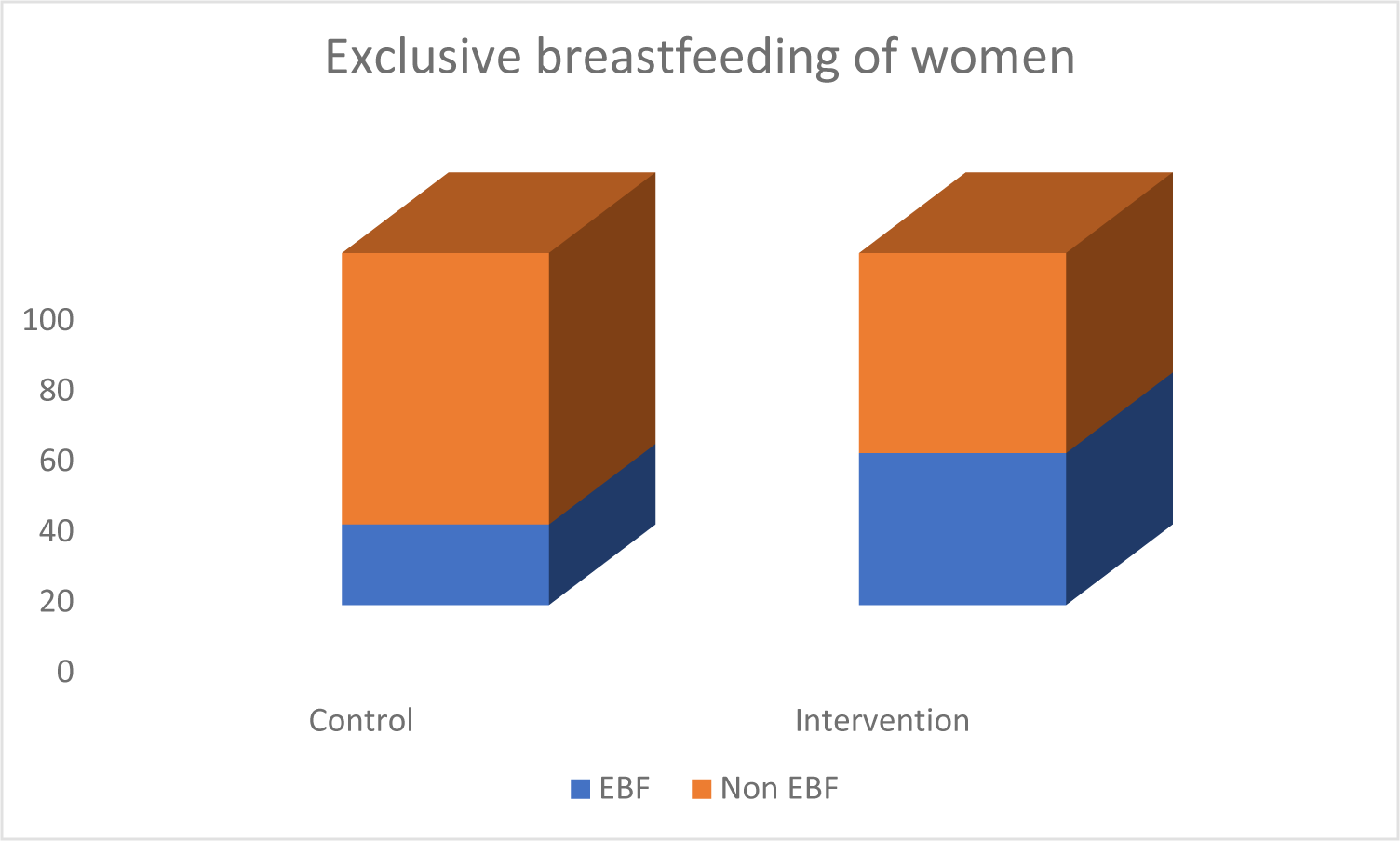
Exclusive breastfeeding practice of women

**Figure 3:**
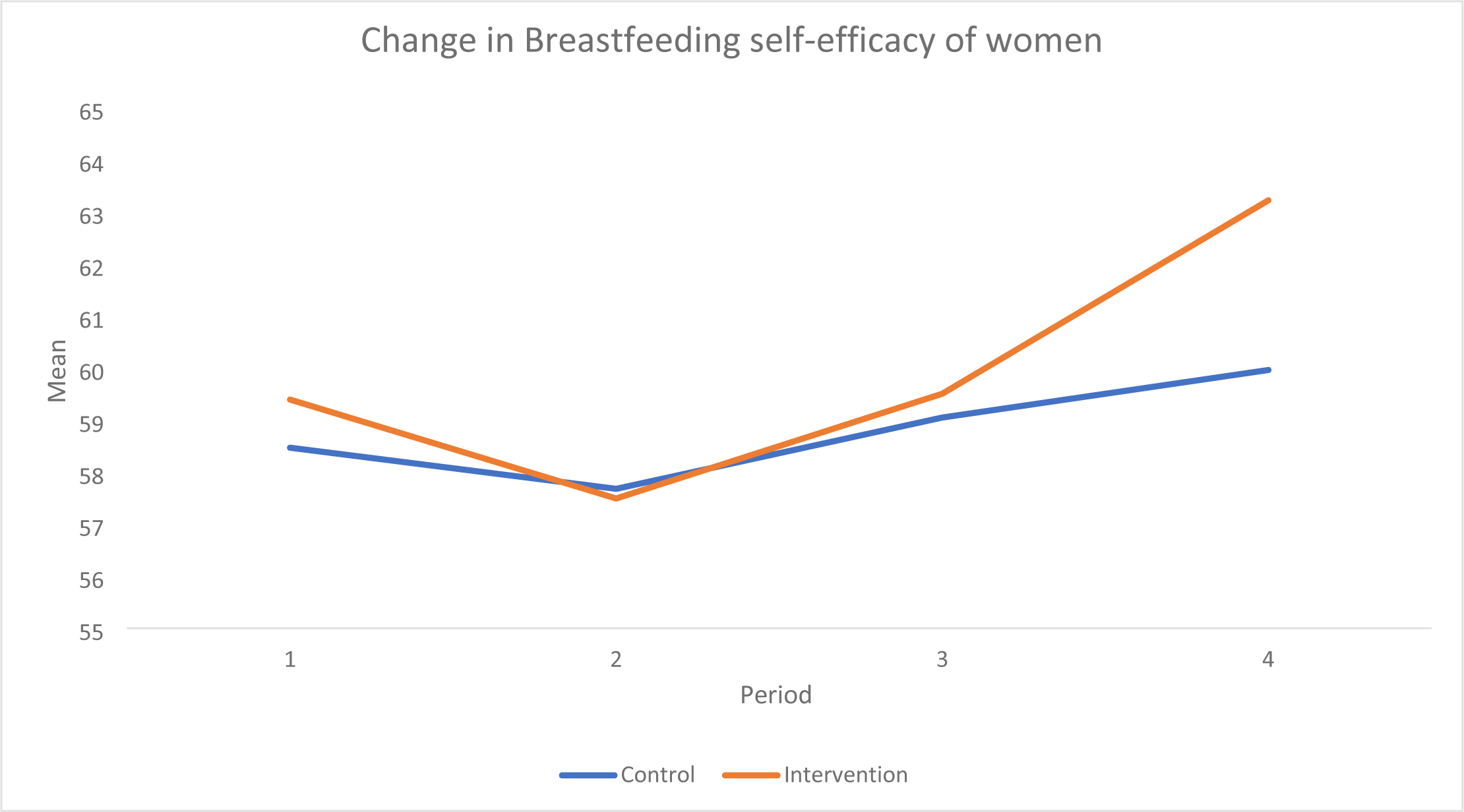
Change in the breastfeeding self-efficacy mean score of women

The figure below explains the trend in the breastfeeding self-efficacy (BFSE) mean score of women from baseline (pre-intervention) to endline (one, three, and six months post-intervention). At baseline, the BFSE mean score was 58.47±5.83 and 59.39±7.52 for control and intervention groups, while it was 57.68±5.58 and 57.49±5.29; 59.05±5.03 and 59.51±5.47; 59.96±6.36 and 63.23±5.59 at one month, three months, and six months postpartum for control and intervention groups, respectively (p<0.05). There was an increase in the mean breastfeeding self-efficacy score of women in the intervention group at three months and six months postpartum as compared to the control group (p<0.001). Age, marital status, delivery mode, and occupation were identified as predictors of breastfeeding self-efficacy.

## DISCUSSION

The findings of the socio-demographic characteristics gave an overall mean age of 27.00±6.20 and 28.40±6.50 for intervention and control groups. Some demographic variables were also identified as predictors of breastfeeding self-efficacy and exclusive breastfeeding practices of the women. Some of these factors include maternal age, marital status, post-secondary education and occupation while for exclusive breastfeeding practices of the women. This is similar to that of Al-Thubaity et al., (2023); Salarvand et al., (2023); Corby et al., (2021) and Economou et al., (2021). The drama intervention had a significant effect on the breastfeeding of women in the intervention group as compared to those in the control group. At three and six months postpartum, women in the intervention group had significantly higher mean breastfeeding self-efficacy scores than women in the control group. However, most of the intervention also initiated breastfeeding early as well as practice exclusive breastfeeding unlike those in the control group. This is consistent with other studies by Almohanna *et al. (2020),* Azimi *et al*. (2020), Arban *et al*. (2018), Dodt *et al.,* 2015; McQueen *et al*., 2011) that evaluated the effect of educational interventions, flipcharts, health talks, counseling, and internet based electronic technology interventions on breastfeeding self-efficacy and breastfeeding outcomes of women.

### Breastfeeding initiation and exclusivity

Majority of the participants (65.5%) in the intervention group started breastfeeding within the recommended WHO standard of one hour after delivery, and 43.2% exclusively breastfed their infants for six months postpartum. This result is consistent with the finding of Ansari et al. (2014). About one third of the infants were fed first breast milk (colostrum) and also had skin-to-skin contact with their mothers. Few challenges were also reported by participants to affect the practice of exclusive breastfeeding and this include: nipple pain, insufficient breastmilk, sunken fontanel (*Oka-ori*), inability to sleep well, eating regularly, and mother-in-law advice on the introduction of herbs and water to infants before six months. These results concur with that of Awaliyah *et al*. (2017), Liu *et al*. (2011), and Bunik *et al*. (2010). Despite this, the drama intervention was observed to be effective in improving the breastfeeding initiation rate and exclusive breastfeeding of participants.

### Breastfeeding self-efficacy

The results from these findings show the breastfeeding self-efficacy mean score at baseline to be higher than 46.4 reported by McQueen et al. (2011), 55.8 by Dennis (2003), and 55.5 by Wutke and Dennis (2007). The variation in the mean score might be due to the difference in location and sample size. At three and six months postpartum, the mean score of women in the intervention group was higher than those in the control group. This is inline with the results of intervention work using workshop, workbooks, pamphlets, videos, movies, flipcharts, peer educators, and telephone (6, 8-9,25-26, 28-32).

The mode of delivery had a significant positive effect on the breastfeeding self-efficacy of the participants. Participants who had spontaneous normal vaginal delivery and marital status had high breastfeeding self-efficacy. This is in line with the work of Ngo *et al*. (2019) and Economou *et al*. (2021), as most women who are married tend to have support from their partners on breastfeeding self-efficacy, thereby enhancing their practice of exclusive breastfeeding.

### Conclusion

There was an increase in the breastfeeding self-efficacy, breastfeeding initiation, and exclusive breastfeeding of women in the intervention group compared to those in the control group from baseline to six months postpartum. The drama intervention was therefore effective in the increase of breastfeeding self-efficacy to achieve the WHO recommended exclusive breastfeeding rate. The practice of exclusive breastfeeding and mothers’ breastfeeding self-efficacy was also found to be influenced by predictors such maternal age, marital status, occupation, and education. To increase the exclusivity rate, policies should also be developed on the use of educational entertainment by healthcare professionals, such as theater.

## Conflict of interest

*This study was sponsored by Nestle Research Foundation, Switzerland*.

## Data Availability

All relevant data are within the manuscript and its supporting information files

## Acknowledgements

I would like to appreciate all the research participants from the selected communities in the two Local Government Areas for their voluntary consent and participation during the period of the study. I would also like to thank the peer support group in the communities who helped to monitor these women from pregnancy to the period of their delivery.

## Notes

### Competing Interest Statement

The authors have declared no competing interest.

### Funding Statement

The award grant was given to Mrs Ogundairo Yetunde Omotola by Nestle Foundation for the study of problems of Nutrition in the world, Lausanne, Switzerland.

